# Network analysis applied to DASS-21: Emergence of a new dimension

**DOI:** 10.1101/2022.07.29.22274386

**Authors:** Marco Antônio Silva Alvarenga, Paulo Felipe Ribeiro Bandeira, Carollina Souza Guilhermino, Tiago Geraldo de Azevedo, Kelly Fernandes Olímpio, Camila Kersul, Glacithane Lins da Cunha, Juliana Alves-Teodoro, Pricila Cristina Correa Ribeiro, Marcela Mansur-Alves, Maycoln Lêoni Martins Teodoro

## Abstract

The DASS-21 has been studied in different samples and cultures as a brief tool for screening and referral to specialized interventions, thus presenting the prerogative to be characterized as a complex system (CS). CS is a new approach to data analysis assumes that items on a scale are components integrated as a network. Despite this, to date network analysis has not been applied to verify the psychometric properties of the DASS-21. This was a non-clinical sample consisting of college students and professionals (N = 4017), aged 18 years or older (M = 31.16; SD = 10.308), from different Brazilian regions. The data collection was done through electronic forms composed by a sociodemographic questionnaire and DASS-21. Participants could forward the form to other potential respondents. Uni, bi, and multivariate analyses were used, and, among them, exploratory graph analysis (EGA) and boostrap EGA (BootEGA). EGA e BootEGA generated a model with four factors. The four-factor model from DASS-21 showed better fit rates compared to the others replicated in this research. The new four factor model has excellent composite reliability and is invariant regard to gender and type of activity performed. This research was composed of a non-probabilistic and convenience sample, without equitable geographical distribution of the participants and whose answers to this study were provided only by the web-based forms. The DASS-21 presented a new factor model composed of four distinct dimensions with excellent intrinsic features.

## 1. Introduction

The short form of the Depression, Anxiety and Stress Scale (DASS-21) is a widely used self-report instrument to assess affective-emotional changes related to mood and anxiety [1–3]. This version presents a model with good discrimination among symptoms related to depression, anxiety and stress in clinical and non-clinical samples [4–6] comprised of adolescents [7,8] college students [9,10] healthcare professionals [11] psychoactive substance users [12] and athletes [13]. The depression subscale features items related to moodiness, hopelessness and lack of motivation; the anxiety subscale reports response promptness and physical changes; and the stress subscale refers to persistent tension, irritability and a poor tolerance for frustration [14].

As of the time of writing this article, DASS-21 has been adapted for at least 55 different linguistic-cultural contexts across all continents [15]. There is research on this scale in Oceania [16], Africa [17–19], America [14,20–24] and Eurasia [25–36]. Thus, DASS-21 has become a widely used instrument to assess the severity of burden and affective changes. Despite the consistency of the results found in the investigations carried out, there is divergence about the dimensional structure of this instrument.

Exploratory factor analysis (EFA) and confirmatory factor analysis (CFA) have been the methods most used to verify the psychometric properties of the scale [2], often indicating a three-factor model. However, other models for DASS-21 have been reported from such analyses. Henry and Crawford [5], for example, have shown a model expressed by three factors (represented by items from the depression, anxiety and stress subscales) and also by a single factor (general psychological distress), calling it a bifactor model. Uni- and bidimensional models are also described in the literature: some items from the depression and anxiety subscales are taken to form just one dimension [37] the depression and anxiety subscales are classed as one dimension and the stress subscale as another [38] the depression and stress subscales are classed as one dimension and the anxiety subscale as another [23]; the anxiety and stress subscales are classed as one dimension and the depression subscale as another [39,40] and a bidimensional model expressed by two dimensions distinguished from those previously mentioned – physiological arousal and generalized negativity [41].

Some recent studies have employed other methods for analyzing DASS-21, namely the exploratory structural equation model (ESEM: [16,42] and the bifactor ESEM (BESEM: [2]. ESEM has the advantage of combining EFA and CFA in such a way that the dimensions described are not related to each other [43]. Using ESEM, both Johnson et al. [16] and Kyriazos et al. [42] evidenced a three-dimensional model for DASS-21 versions with different numbers of items. BESEM makes it possible to combine two simultaneous analysis methods for a bifactor approach [44]. However, Gomez et al. [2]. demonstrated that a general factor model was more satisfactory when they applied this method to DASS-21.

Other analyses provide evidence for maintaining the dimensional structure of the scale across different groups, such as the factor invariance calculation [45,46]. This technique has been applied to DASS-21 and has shown factor invariance for the three-dimensional model across a diverse group of college students [47]: from different countries [24,48], gender [29,49], age groups [35,50] and for the full and nine-item versions of the scale for a non-clinical sample [42]. Other studies have reported invariance for the bifactor model between clinical and non-clinical samples [51] and among different countries [52].

Despite the variety of methods applied to DASS-21, this instrument has not yet undergone an analysis based on complex systems and network science. In several areas, phenomena are investigated as systems that, owing to their sensitivity to initial conditions, non-linearity, emergent behavior and self-organization, can be classified as complex [53]. Psychometric research has presented this perspective in different fields of study in recent years, such as in psychopathology [54], persistent somatic symptoms [55], major depression [56] and anxiety and depression symptoms [57]. The network perspective investigates the multivariate data structure of DASS-21 in different situations, for example, by investigating associations between variables, identifying more important and intervention-sensitive variables from centrality indices and exploring the dimensional structure of the psychological scales [58].

Exploratory graph analysis (EGA) and bootstrap EGA (BootEGA) are two different types of network analysis. EGA and BootEGA were developed to estimate the number of dimensions in multivariate data using non-directional network models [59,60]. These analyses are part of a new perspective known as “network psychometrics”. Under this approach, attributes are conceptualized as causally linked variables (observable) that form an emergent network pattern or topology. This means that the organization of items is characterized as a complex system according to contextual variables [58]. Considering all the above, this research aims to employ contemporary (EGA and BootEGA) and traditional (CFA, composite reliability and factor invariance) analyses to DASS-21.

## 2. Methods

### 2.1. Study design

This is a descriptive, correlational and quantitative research analysis because it represents the characteristics of the sample without having control over the variables described, understanding the correlations (whether linear or in a complex system) among those variables and establishing parameters for comparison from the observations that have been made [61]. It is a cross-sectional study employing the snowball technique to increase the sample size [62], with a non-probability sample type since there was no randomization for the selection of participants [63].

### 2.2. Participants

The minimum sample size was calculated using G*Power 3.1 software for iOS [64], adopting the parameter test of variance equality (two-sample case: ratio variance1/variance0 = 1.5) because usually there are twice as many females as males in studies involving DASS-21 (see 40,64,65). We further adopted the recommendations of a sample consisting of 10 respondents per item but 25 for more conservative analyses and for CFA [45] and EGA [59]. The minimum sample comprised 714 participants, 416 of which were females.

Participants included were 18 years of age or older and a current student or professional from any of the five Brazilian regions. We excluded the following: incidental responses from different profiles intended in this research; participants who did not complete the DASS-21; and self-reported psychiatric and/or neurological diagnosis in the last two months from the date of response.

This study comprised 4017 people with a mean age of 31.16 years (SD = 10.308). Most participants were female (N = 2791, 60.5%), college students (70.3%), self-described as Caucasian (60.8%) or brown (29.1%), single (63.0%) and with a monthly household income of 2–10 minimum wages (58.6%). Many participants did not disclose which Brazilian region they lived in (70.4%); however, among the respondents, most reported living in the Southeast region (76.4%). The mean of the DASS-21 total score was 23.98 (SD = 15.33) for the overall sample, with a significant mean difference for all items and for the DASS-21 total score between genders, with minimum to moderate effect size (see Table 1).

**Table 1.**
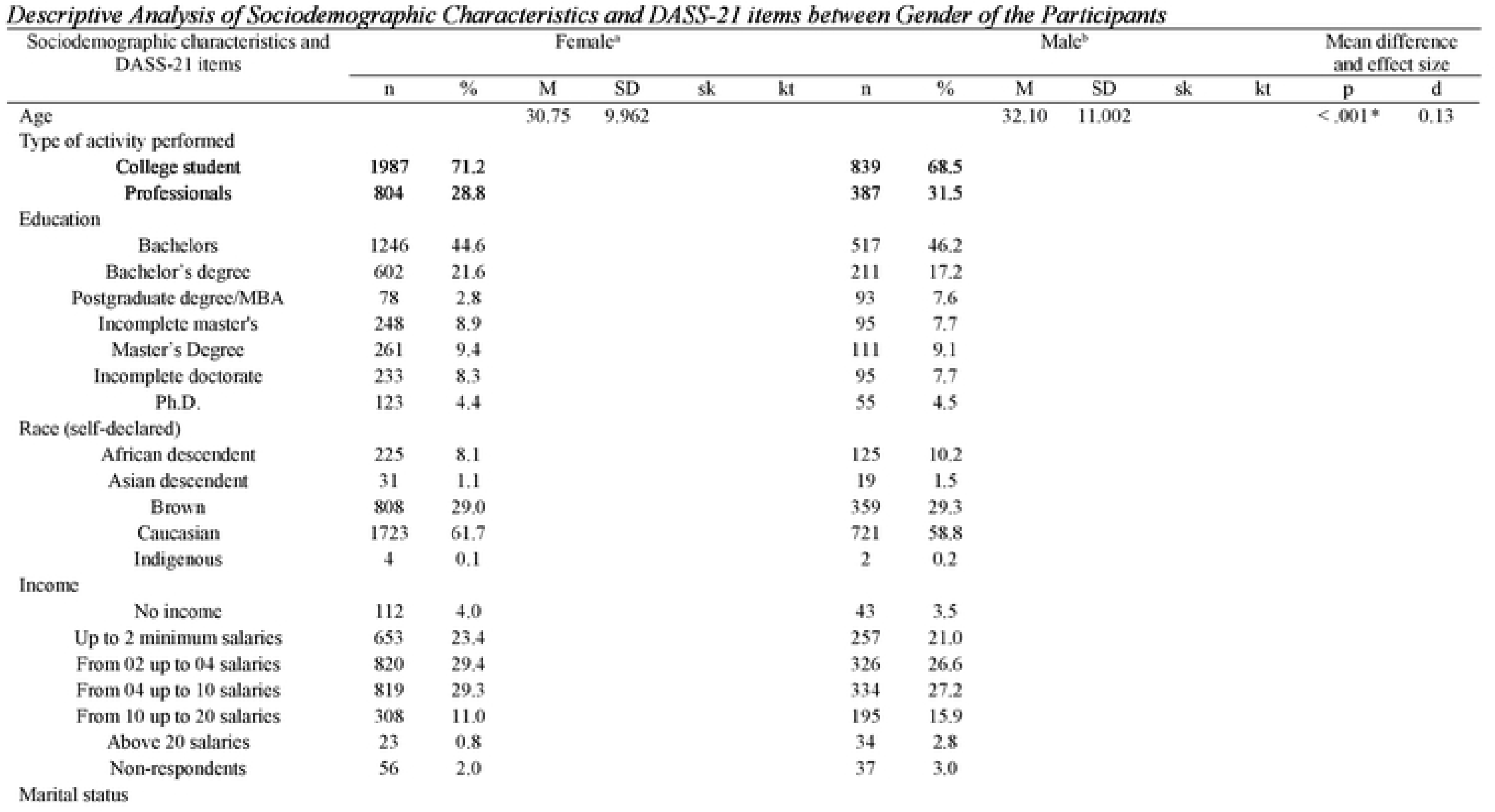
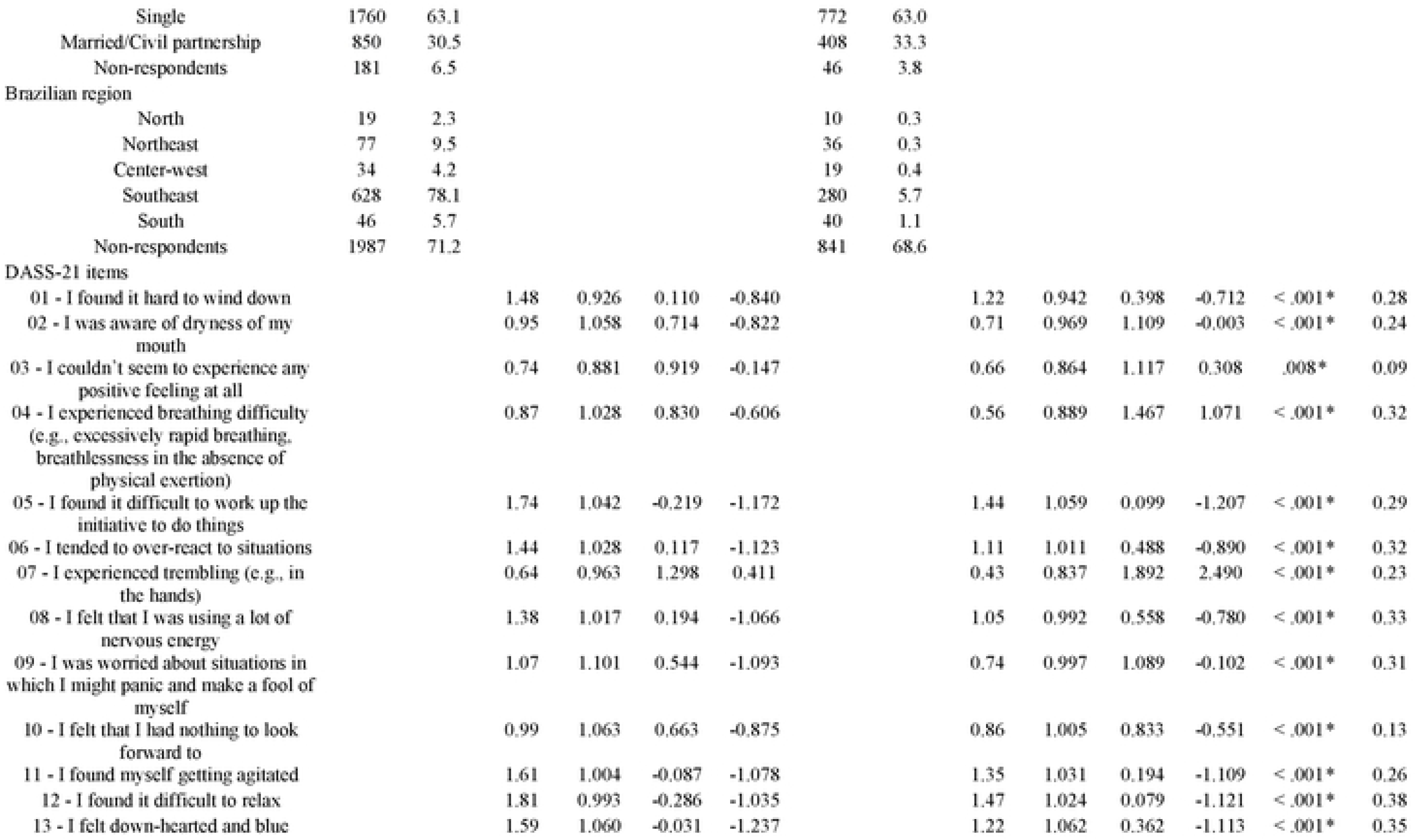
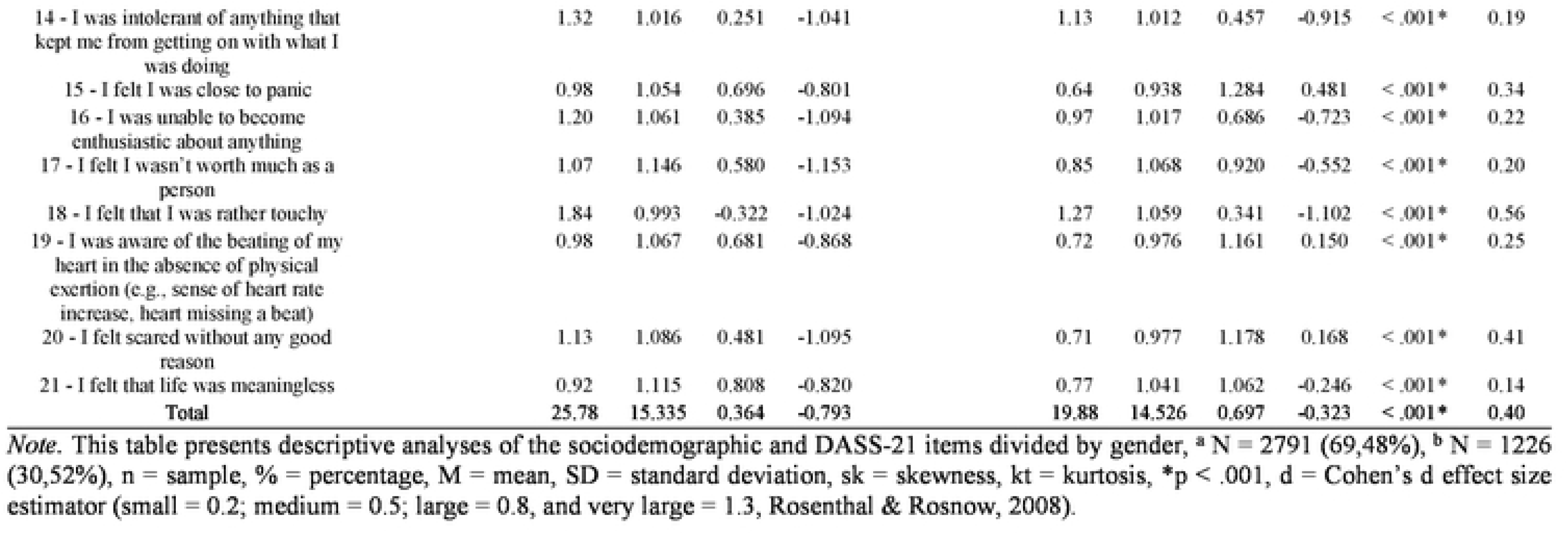
Descriptive Analysis of Sociodemographic Characteristics and DASS-21 items between Gender of the Participants.

### 2.3. Measures

A sociodemographic questionnaire was developed for the purpose of obtaining data on age, gender (female, male, intersex), marital status (single, married, divorced, etc.), self-reported race (African descendant, Asian descendant, brown, indigenous and white), school background (elementary school, high school, college, etc.), family income, occupation (college student, professional) and region of residence (North, Northeast, Center-West, Southeast, South).

DASS-21 [3,68] was adapted to the Brazilian population by Vignola and Tucci [69]. It is composed of 21 items, with seven items for each subscale: depression (α = .92), anxiety (α = .86) and stress (α = .90). The items are rated on a four-point Likert scale ranging from 0 (“did not apply at all”) to 3 (“applied a lot or most of the time”).

### 2.4. Procedures

This investigation was carried out exclusively by web-based data collection (due to the COVID-19 pandemic) from April to August 2020.

The research team prepared an invitation letter explaining the purpose of the study, with a link providing access to the online form comprising the consent form, the sociodemographic questionnaire and DASS-21. It was forwarded by email to universities, companies/managers, hospitals and health/psychosocial care centers in different regions of the country and was also published on Facebook and Instagram accounts. Participants could also forward the form to other potential contributors.

This study was approved by an ethics and research committee involving human subjects under CAAE registration code 07077019.3.0000.5149, according to National Health Council (CNS) resolution numbers 466/2012 and 510/2016.

### 2.5. Statistical and data analysis

#### 2.5.1. Descriptive analysis

Descriptive analyses were used to describe the characteristics of the sample [70]. Skewness, kurtosis, mean difference and effect size tests were applied to continuous variables, with gender as a comparison factor. When pertinent to the analyses, a p value of < .05 was adopted.

#### 2.5.2. EGA

EGA is a recently developed method to estimate the number of dimensions in multivariate data using undirected network models [59,60]. EGA first applies a network estimation method followed by a community detection algorithm for weighted networks [71].

##### 2.5.2.1. Network estimation method

This study applied the graphical Least Absolute Shrinkage and Selection Operator (GLASSO [72,73], which estimates a Gaussian graphical model (GGM [74]) where nodes (circles) represent variables and edges (lines) represent the conditional dependence (or partial correlations) between nodes, given all other nodes in the network.

LASSO uses a parameter called lambda (λλ), which controls the sparsity of the network. Lower values of λλ remove fewer edges, increasing the possibility of including spurious correlations, whereas larger values of λλ remove more edges, increasing the possibility of removing relevant edges. When λλ = 0, the estimates are equal to the ordinary least-squares solution for the partial correlation matrix. In this study, the ratio of the minimum and maximum λλ was set to .1 (.5 is the higher value).

The popular approach in the network psychometrics literature is to compute models across several values of λλ (usually 100) and to select the model that minimizes the extended Bayesian information criterion (EBIC [75,76]. The EBIC model selection uses a hyperparameter gamma (γγ) to control how much it prefers simpler models (i.e., models with fewer edges [77]. Larger γγ values lead to simpler models, whereas smaller γγ values lead to denser models. When γγ = 0, the EBIC is equal to the Bayesian information criterion. In this study, γγ was set to 0. In the network psychometrics literature, this approach has been termed EBICglasso and is applied using the qgraph package.

##### 2.5.2.2. Community detection algorithm

The Walktrap algorithm [78] is a commonly applied community detection algorithm in the network psychometrics literature [59,60]. The algorithm begins by computing a transition matrix where each element represents the probability of one node traversing to another (based on node strength or the sum of the connections to each node). Random walks are then initiated for a certain number of steps (e.g., 4) using the transition matrix for probable destinations. Using Ward’s agglomerative clustering approach [79], each node starts as its own cluster and merges with adjacent clusters (based on squared distances between each cluster) in a way that minimizes the sum of squared distances between other clusters. Modularity [80] is then used to determine the optimal partition of clusters (i.e., communities). The Walktrap algorithm was implemented using the igraph package in R [81].

#### 2.5.3. CFA and internal consistency

CFA using the unweighted least-squares estimator was conducted to test the internal structure of DASS-21 in seven different models, including the model originated by EGA. An adequate fit was considered when comparative fit index (CFI) and Tucker-Lewis index (TLI) values were > .90; values of > .95 indicated a good fit [82]. Root-mean-square error of approximation (RMSEA) values of .08 and .06 indicated an acceptable fit, with values < .05 indicating a good fit [82]. Factor loadings were considered adequate if they were ≥ .40. Internal consistency was calculated from composite reliability [83]. Indices with values of ≥ .80 were considered adequate [84].

#### 2.5.4. Factor invariance

Factor invariance of CFA for gender and occupation was tested on a DASS-21 four-factor model: configural invariance (equality for fit); weak or metric invariance (equality for factor loadings); strong invariance (equality for item interceptions); and strict invariance (equality for residual variances or uniqueness) [46]. The fit of the configural model data was measured using CFI, TLI and RMSEA. The configural model would be rejected if it displayed CFI < .90 or RMSEA ≥ .08. The weak, strong and strict invariance models would be rejected if they displayed ΔCFI > .005 and ΔTLI > .005.

The database for this research can be downloaded and cited from the following link https://osf.io/xpabk/.

## 3. Results

### 3.1. EGA and BootEGA

Figure 1a provides the estimated dimensionality of DASS-21 obtained using EGA. Four dimensions were estimated, with items distributed as follows: 01, 06, 08, 11, 12, 14 and 18 for the stress dimension and 03, 05, 10, 13, 16, 17 and 21 for the depression dimension; the anxiety dimension was divided into two, one with items 02, 04, 07 and 09 and the other with items 09, 15 and 20. Figure 1b provides the median dimensionality structure of the network (i.e., the network calculated as the median of the estimated partial regularized correlations between bootstrap networks).

**Figure 1.**
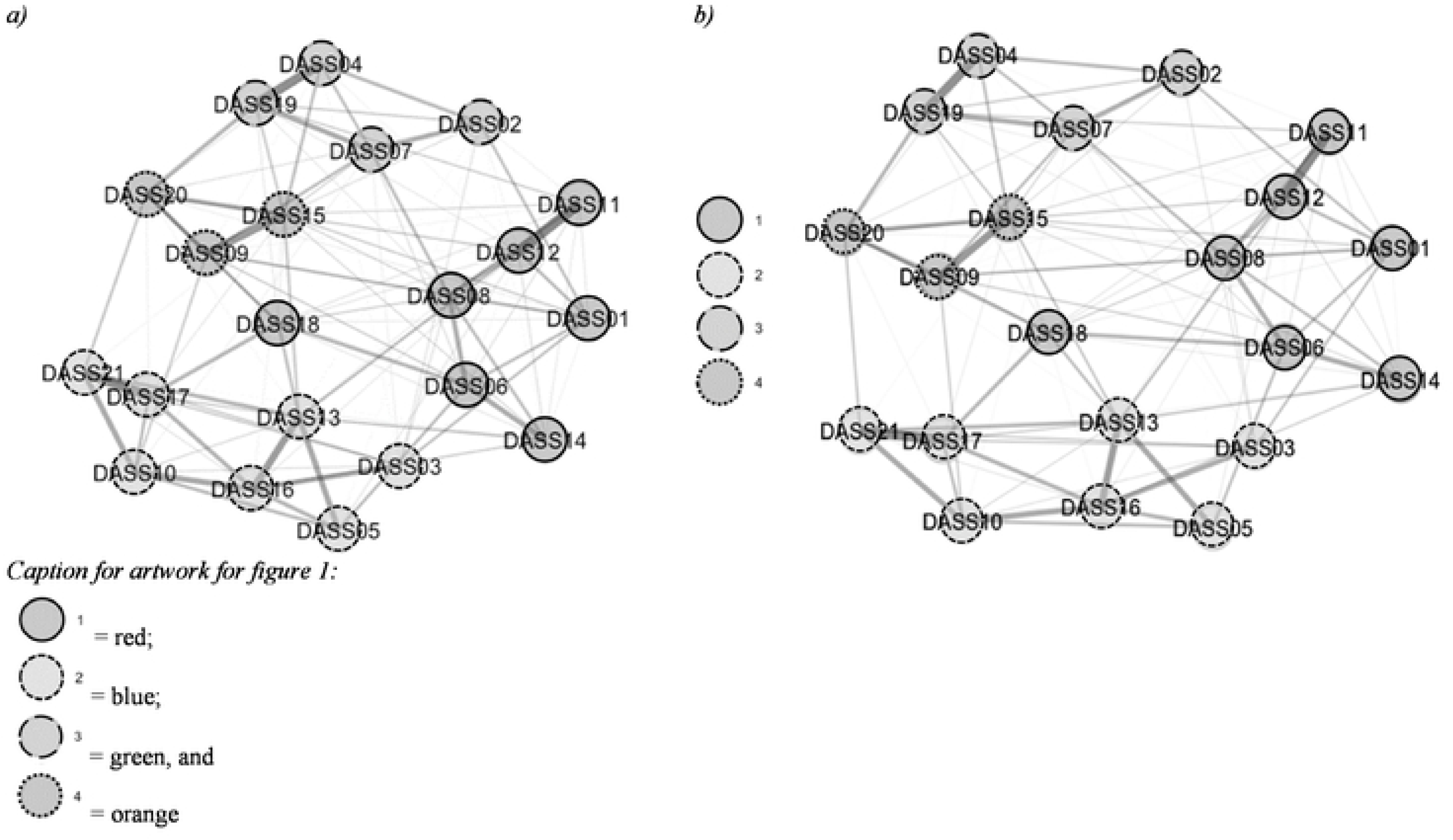
Dimensionality structure of DASS-21 from EGA (a) and BootEGA (b). *Note.* Representation of the dimensional structure of the new DASS-21 model from EGA (a) and BootEGA; The lines in both figures and their thickness express the relationship between items. Items that have the same color are represented as belonging to the same dimension; red (1) = stress items, blue (2) = depression items, green (3) = anxiety items, and orange (4) = panic items. BootEGA supported the 4-factor model by 72.6%.

The original EGA structure with four dimensions was found in most of the dimensional solutions estimated via BootEGA. In addition to the four-factor dimensional structure in 72.6% of the interactions, a three-dimensional structure was observed in 26.5% and a five-dimensional structure in 9% of interactions. Item replication analyses indicated good replicability across all items in the four-dimensional model (> .65) (Figure 2).

**Figure 2.**
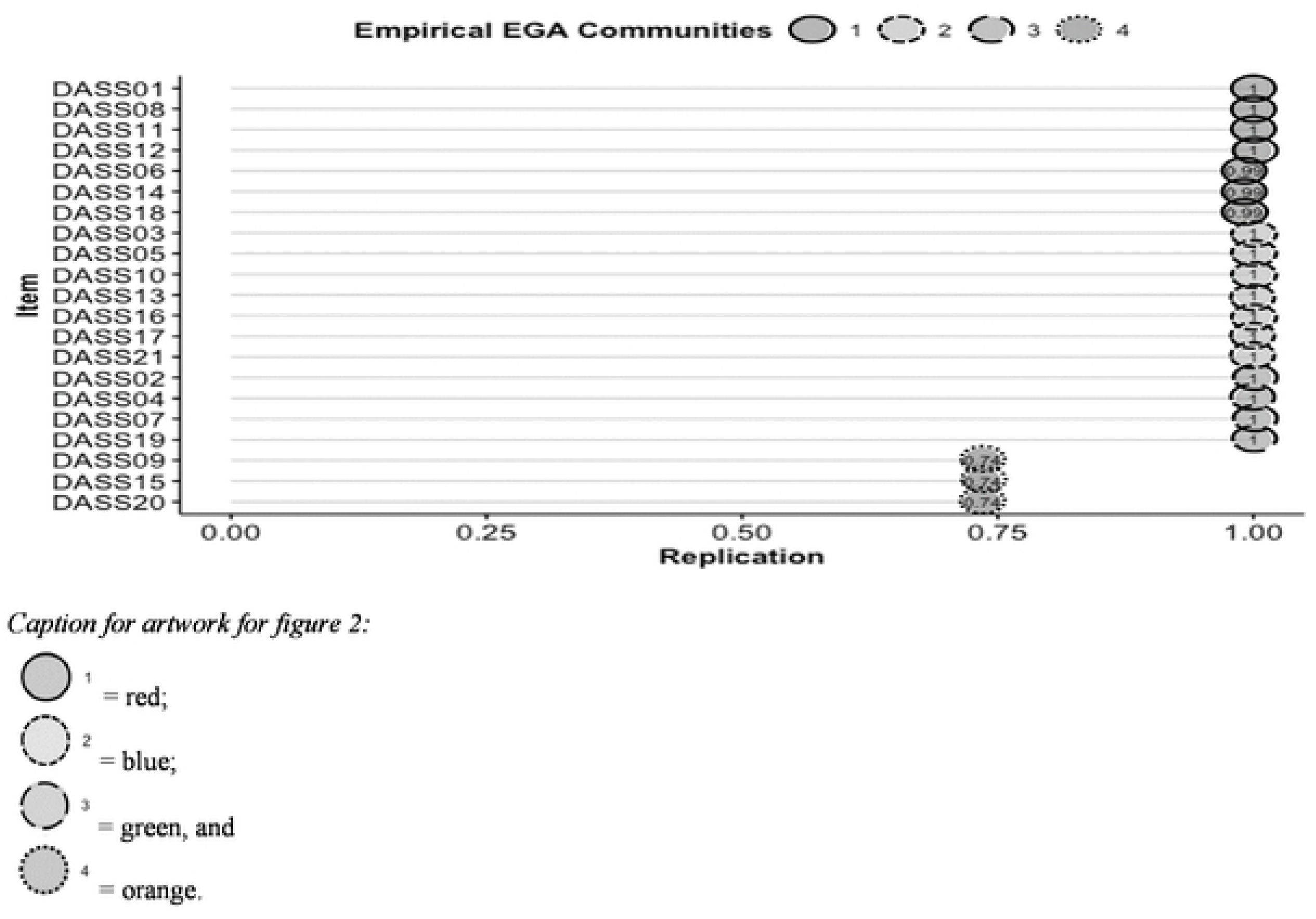
Replication of the DASS-21 items for each of the original EGA dimensions. *Note.* Replication of EGA DASS-21 Model for four dimensions. Best fits of replication are > .65.

### 3.2. CFA and internal consistency

The factor loadings of the DASS-21 items revealed appropriate factor loadings for all seven models tested (see Table 2).

**Table 2.**
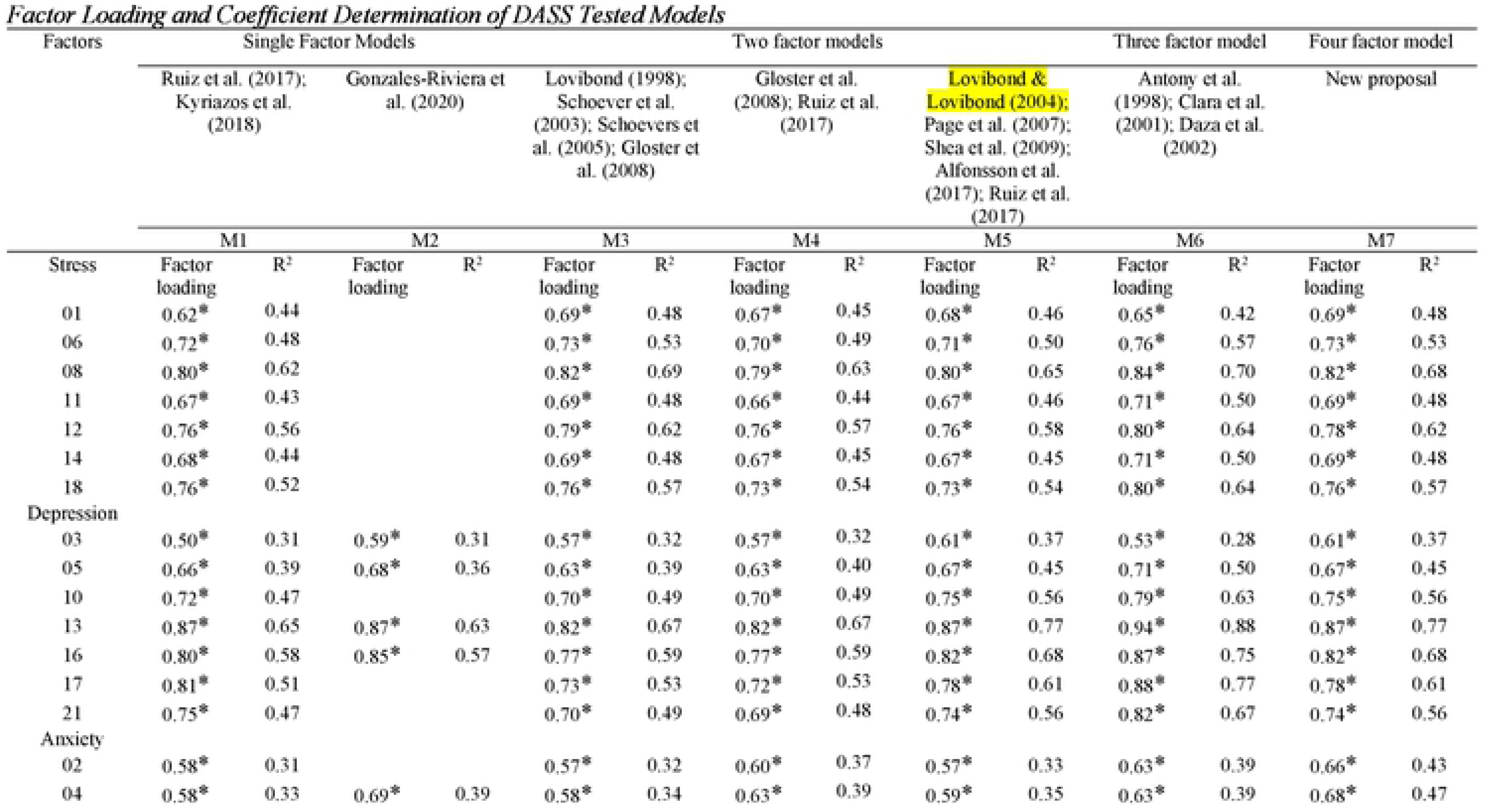
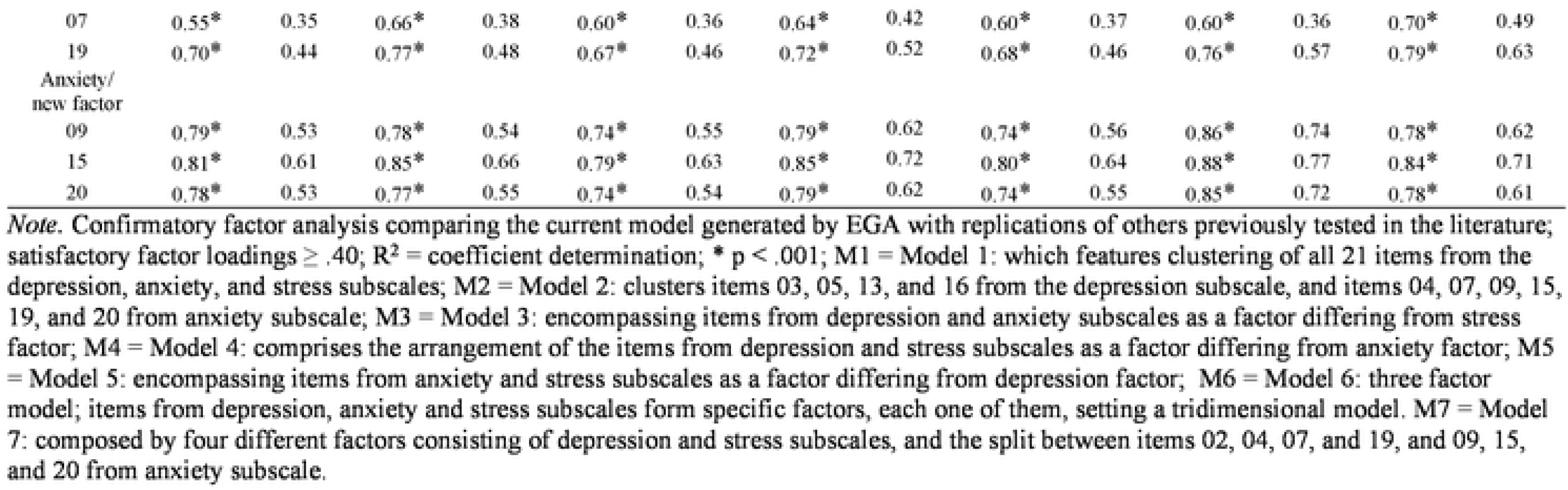
Factor Loading and Coefficient Determination of DASS Tested Models.

All models showed very good fit indices (CFI and TLI > .95 and RMSEA ≤ .08) but Model 7 was the best of all because it also showed a non-significant chi-square value (see Table 3).

**Table 3.** Fit Indexes of Replicated and New DASS-21 Models.

| Model | $\chi^2$ | df | CFI | TLI | RMSEA | 90% CI RMSEA | SRMR |
| --- | --- | --- | --- | --- | --- | --- | --- |
| M1 | 2954.93* | 189 | 0.99 | 0.98 | 0.060 | 0.040-0.070 | 0.060 |
| M2 | 1186.33* | 35 | 0.98 | 0.97 | 0.080 | 0.080-0.090 | 0.070 |
| M3 | 2451.094* | 188 | 0.98 | 0.98 | 0.055 | 0.053-0.057 | 0.074 |
| M4 | 2221.741* | 188 | 0.98 | 0.98 | 0.052 | 0.050-0.054 | 0.050 |
| M5 | 1564.710* | 188 | 0.99 | 0.99 | 0.043 | 0.041-0.045 | 0.043 |
| M6 | 1060.12* | 186 | 1.00 | 0.99 | 0.030 | 0.030-0.040 | 0.030 |
| M7 | 899.09 | 183 | 1.00 | 1.00 | 0.030 | 0.029-0.033 | 0.030 |

*Note.* Confirmatory fit indices for each of the tested DASS-21 models, including the one with a new fourth dimension; the name of the models, factors, subscales and comprising items were described in table 2; $\chi^2$ = chi-square; \* p < .01; df = degrees of freedom; CFI = Comparative Fit Index; TLI = Tucker Lewis Index; RMSEA = Root-Mean-Square Error of Approximation; CI RMSEA = Confidence Interval of Root-Mean-Square Error of Approximation; SRMR = Standardized Root Mean Square Residual.

The composite reliability values were .901 for depression, .894 for anxiety, .890 for stress and .842 for the new dimension.

### 3.3. Factor invariance

The overall fit indices were adequate for the configural model and there was invariance in all the procedures applied. These results support measurement invariance for the four-factor model and indicate the same underlying constructs observed for both men and women as for the type of activity performed by participants (see Table 4).

**Table 4.**
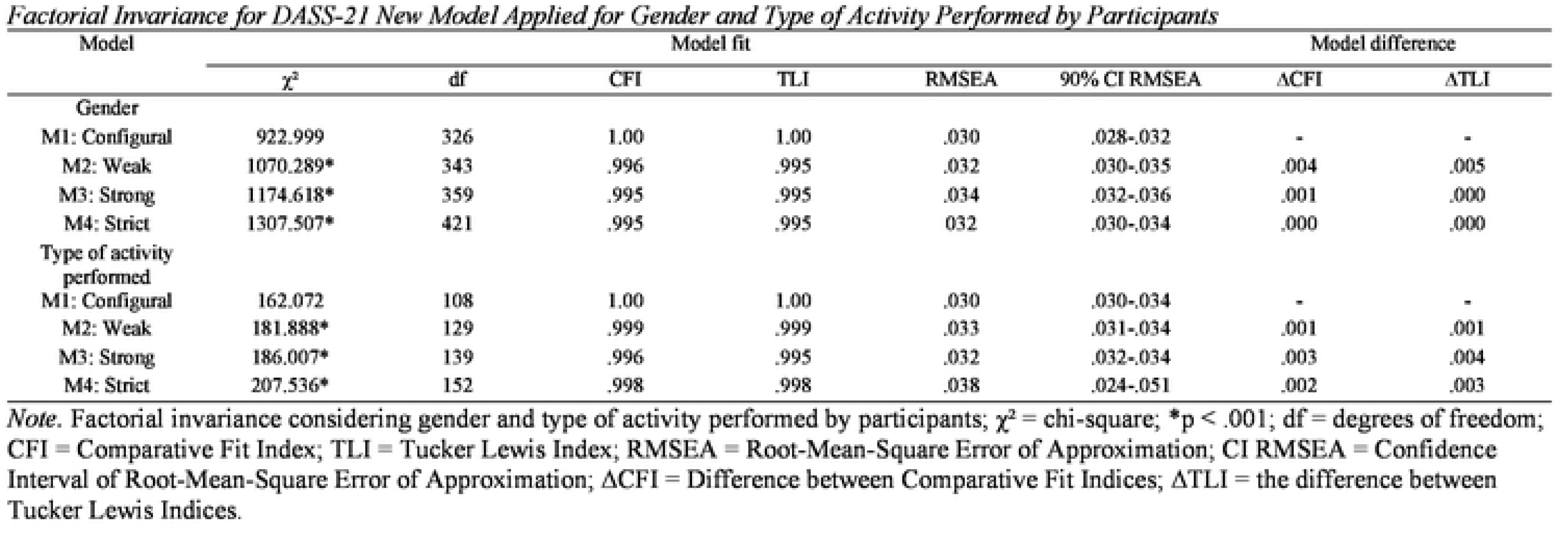
Factorial Invariance for DASS-21 New Model Applied for Gender and Type of Activity Performed by Participants.

## 4. Discussion

The principal purpose of this study was to examine the psychometric properties of DASS-21 from a network psychometrics perspective. The results revealed four dimensions in the scale, instead of three. CFA demonstrated that this model presents excellent fit indices – individual and overall – even better than those replicated from others previously tested in the literature. The composite reliability indices showed excellent values. The dimensions of the new model were invariant between sex and type of activity performed by participants. Thus, the applicability of the dimensional scores of the scale was demonstrated.

### 4.1. Sample characteristics and new approach for psychometric properties of DASS-21

It is common for studies to point out significant differences in gender between the subscales of DASS-21 [20,91], showing a higher prevalence of negative affective experiences for women compared to men [92]. This research reported significant difference between females and males’ participants for each item of the scale and the total score, with small to medium effect sizes, which corroborates data already established by the literature.

Research regarding the psychometric properties of DASS-21 has mostly used EFA and CFA as the main tools to check the association between the scale items. We expect to see a greater amount of these types of studies due to the extensive use and adaptation of DASS-21 for different cultural contexts [2,23,37]. However, DASS-21 showed unsatisfactory confirmatory fit indices, even with a three-dimensional model being the most promising, as in the studies by Brown et al. [1], Clara et al. [90], and Daza et al. [66]. Other research also reported problems regarding the factor configuration of the DASS-21, since confirmatory models displayed appropriate fits on deleting one or more items or on associating residual variances between them [see 5,8,17,37,39,48,51,93].

Unsatisfactory fit indices for DASS-21 dimensions persevered even when using ESEM and BESEM. It happens due to latent variables being measured through exploratory factor analysis, and in that sense, the explanation about a relationship between latent variables becomes more difficult due to multiple item cross-loadings [94]. Johnson et al. [16] observed that items related to anxiety factor loaded into the two other factors of the scale. Kyriazos et al. [42] showed there were excellent fits for a trifactor model for a shortened version of the scale. However, the relationship between the dimensions was very strong, above .870 in all cases. Gomez et al. [2] verified that some items from both the anxiety and the stress subscales were poorly defined in terms of their target dimensions. Further studies on DASS-21 based on Item Response Theory (IRT) have also reported problems of item fitting [38,95,96]. Thus, considering previous outcomes in the literature and the complex nature of the groups and scenarios that have been observed, then, we decided to employ EGA and BootEGA. In addition, a recent review of the literature has not yet reported studies in which such methodology has been applied to DASS-21.

Network analysis presented in this paper allows observing the strength of association among the items in which it is possible to verify the formation of clusters [59] and therefore does not aim to recognize a common latent trait [58]. Furthermore, using EGA as a technique to process the data, it was possible to identify four factors instead of three. The depression and stress factors were kept, while the anxiety subscale was divided into items 02 (I was aware of dryness of my mouth), 04 (I experienced breathing difficulty), 07 (I experienced trembling), and 19 (I was aware of the beating of my heart in the absence of physical exertion) in one factor, and items 09 (I was worried about situations in which I might panic and make a fool of myself), 15 (I felt I was close to panic), and 20 (I felt scared without any good reason) in another. Items from anxiety subscale have previously presented problems regarding their theoretical dimension [37], as they have been distributed among others [2,41 97]. However, this configuration among the anxiety subscale items had not yet been reported. In addition to the one new dimension, the values of empirical EGA communities were greater than .65 across items and their respective dimensions (see Figure 2). BootEGA checked the consistency of the four-dimensional model derived for DASS-21. In addition, it was possible to establish the topological similarity between EGA and BootEGA from visual analysis (see Figure 1a and b).

CFA applied to this new model revealed better individual and overall fit indices, including a non-significant chi-square, compared to the others tested in this research. No associations were made among the measurement errors of the items and residuals to enhance the tested models. This was justified because all the models presented satisfactory fit indices and to avoid interfere with the parsimony approach in the interpretation of the results [45]. Most research used Cronbach’s alpha to estimate dimensional and global internal consistency of DASS-21, with a few exceptions [16,38,42]. We chose to use composite reliability after the new model was defined, since it considers measurement errors, providing a more robust index to comprise the portion of estimating each item to its dimension [84], whereas alpha is calculated under the assumption that each item equally affects the variance of a factor [98,99]. The four factors showed excellent internal consistency.

Previous studies have reported evidence for factor invariance for DASS-21 three-dimensional model [24,47–49]. The outcomes of this investigation pointed out there is factor invariance for DASS-21 four-dimensional model regarding gender and type of activity performed by participants which enables the scale for comparative statistics proposes. Future studies focusing on this approach can test the invariance of this new model considering other groups.

Both traditional and network psychometrics analyses have evidenced a new version for DASS-21 with four distinct dimensions. However, we have another two tasks: define how the new arrangement it should be displayed and examine which further evidence is available to support the reallocation among the anxiety items scale into two new dimensions.

### 4.2. Other support for a new dimension of DASS-21

The model with three distinct DASS-21 dimensions is the most reported, although the results of this research demonstrated a split among items belonging to the anxiety subscale. Items 02, 04, 07 and 19 remained labeled as anxiety but what would be the most appropriate name for the dimension comprised of items 09, 15 and 20? DASS-21 was developed based on the recognition of symptoms related to affective and anxiety disturbances and works as a clinical screening method, although with no diagnostic intent [68,100–102]. The new dimension was hereby nominated as “Panic” due to its items being related to panic disorder (PD), as a counterpart to generalized anxiety disorder (GAD) referring to the remaining items.

Studies have exhibited similarities between the etiology of PD and GAD [103,104] but they differ in other aspects. Such a distinction was first drawn in the 3rd edition of the *Diagnostic and Statistical Manual of Mental Disorders* ([DSM-III], APA, [105]) based on the responses assigned to pharmacological treatments for each group [106]. Rapee [107] had identified hyperventilation and subtle onset of PD at an older age than people presenting with GAD. Panic is characterized by a set of sympathetic autonomic responses that culminate in anticipatory seizures, subjective discomfort and fear of losing control and/or dying [108–114], whereas GAD can be defined as a clinical condition held from distortions concerning ambiguous situations that are characterized by uncertain consequences [115]. These two clinical conditions exhibit distinct comorbidities. Ibiloglu and Caykoylu [116] demonstrated that PD has greater comorbidity with bipolar disorder, whereas [80] and Wittchen et al. [117] reported greater comorbidity for GAD with depression, dysthymia, post-traumatic stress and social phobia. There are, however, other ways to distinguish PD and GAD besides phenomenological observations [113], such as neurophysiological, cognitive and psychological treatment plans.

People with PD are more sensitive to bodily sensations and prone to increased basal arousal throughout the day than those with GAD or non-anxious people [118]. A higher heart rate previously has been reported for PD when compared to GAD [119]. Analyses using machine learning have demonstrated that heart rate variability (HRV) measurement can differentiate PD from other anxiety disorders [113] and that people with PD are more prone to heart disease and death [120]. People with PD showed higher levels of interleukin 6 (IL-6) compared to GAD and therefore show better treatment response to escitalopram [121]. There is no clear distinction between prefrontal cortex function for emotion regulation in PD and GAD [122] but there is a distinction for several areas related to the emotional processing, social, planning and threat responses [123]. Thus, information processing may be impaired in both PD and GAD and simultaneously differentiates them in terms of cognitive function. Young adults with PD performed lower on verbal fluency compared to those with GAD, for which performance on memory tasks was shown to be impaired [124].

Interventions based on cognitive therapy have been reported to be effective in reducing both panic and anxiety symptoms [125] but varying in the treatment approach [126]. One approach would be psychoeducation, a method in which the problems and their idiosyncrasies are explained [127,128]. Psychosocial interventions for panic symptoms are reliant on exposure, reassessment of physical symptoms and relaxation but are grounded in behavioral changes, re-evaluation of anxiety-inducing situations and cognitive restructuring for anxiety symptoms [127,129]. This arises due to physical responses being misinterpreted and leading to a panic attack for those with PD [130] and by identifying and reinterpreting specific triggers linked to warning in ambiguous situations for those with GAD [131]. Applied relaxation reduces seizures and feelings of loss of control in panic [132] and levels of worry, physical responses and negative thoughts in anxiety [133]. Furthermore, follow-up studies revealed that people with GAD maintained their improved status for at least 12 months after treatment completion, whereas those with PD did not [134].

All the evidence above shows that there is differentiation between panic and anxiety. This research presented the formation of a new cluster from some items on the anxiety subscale. Analysis of the psychometric properties of DASS-21 revealed a distinction between characteristics that were once attributed only to anxiety but are better applied to panic, as described in the literature on PD and GAD. Thus, the results found in this research corroborate that the experience that differentiates panic and anxiety can be identified by DASS-21. In this sense, clinical practices and research that need to differentiate between these two conditions may rely on the use of a brief screening tool.

## 5. Strengths, limitations and implications

The results of this research come from an emerging data analysis methodology [53,58,60]. Network psychometrics constitutes a new paradigm for understanding how items related to a specific construct associate as a complex system [135,136]. Computation of composite reliability, comparison among DASS-21 models already tested in the literature with the current one and observation of factor invariance by gender and type of activity of the participants revealed that the new factor should be on the agenda of upcoming research involving DASS-21, either by verifying its psychometric properties or for correlational studies. In addition, there is other strong empirical evidence supporting the new “Panic” dimension.

In contrast, this study has clear limitations. It is a non-probability and convenience sample where it was not possible to control the distribution of the number of participants among the different regions of the country. Data collection was conducted virtually due to the COVID-19 pandemic and it was not possible, in accordance with social distance protocols, to carry out any part of the research in a face-to-face format.

In future studies, we suggest, initially, the use of classical and contemporary methods with the factor configuration found in this study for clinical samples. Finally, we propose an opportunity to revise the name of the scale. It would be pertinent to call it the Depression, Anxiety, Stress and Panic Scale (DASP-21). However, we believe that the scientific community will decide on the applicability of this name as several different versions of DASS-21 have emerged [see 4,12, 37, 42] even though none has yet described a fourth factor.

## 6. Conclusion

The results of this research have demonstrated the existence of four factors for DASS-21 in a non-clinical sample and this new model has shown improved fit indices. This study highlights the importance of network psychometrics as an analytical method applied to understanding how the items of a scale are arranged. Furthermore, this new factor configuration may support clinical screenings and treatment programs.

## Data Availability

https://osf.io/xpabk/

## Acknowledgements

National Council for Scientific and Technological Development (CNPq), Brazil. We would like to thank Professor Tatiana Cury Pollo, Ph.D. (Federal University of São João del-Rei, UFSJ) for proofreading and contributions to the improvement of this manuscript.

